# Patients’ research priorities and participation in primary ciliary dyskinesia research

**DOI:** 10.1101/2025.04.04.25325266

**Authors:** Yin Ting Lam, Laura Behan, Katie Dexter, Lucy Dixon, Claudia E Kuehni, Leonie Daria Schreck, Jane S Lucas, Myrofora Goutaki

**Affiliations:** Institute of Social and Preventive Medicine, University of Bern, Bern, Switzerland; Division of Respiratory Medicine, University Children’s Hospital Zurich, Zurich, Switzerland; School of Clinical and Experimental Medicine, Faculty of Medicine, University of Southampton, Southampton, UK; PCD Support UK, London, United Kingdom; Division of Paediatric Respiratory Medicine and Allergology, Department of Paediatrics, Inselspital, Bern University Hospital, University of Bern, Switzerland

**Author notes:** **Correspondence:** Myrofora Goutaki, Institute of Social and Preventive Medicine, University of Bern, Mittelstrasse 43, 3012 Bern, Switzerland.

**Keywords:** Rare lung diseases, survey and questionnaires, systemic disease and lungs, patient outcome assessment

## Abstract

**Introduction:** People living with chronic diseases can provide a unique perspective for research that often differs from that of healthcare professionals. This is particularly important in rare diseases like primary ciliary dyskinesia (PCD), with many knowledge gaps and limited research resources. We aimed to assess participation of patients and caregivers in PCD research and identify their research priorities in a mixed-method study.

**Methods:** We conducted in-depth, semi-structured interviews with adults and adolescents with PCD, and caregivers of children with PCD. After verbatim transcription and thematic analysis, we developed an anonymous online survey, translated it into 8 languages, and circulated it widely in collaboration with PCD support groups worldwide and the European Lung Foundation.

**Results:** The findings from the interviews identified key areas to be explored further though the survey including: developing treatments for PCD and increasing knowledge about different topics such as mental health, fertility, upper airway problems, treatment burden, and impact of environment and lifestyle. 399 participants completed the online survey from 29 countries with median age 41 (IQR 33–49), 74% were female. 180 participants (45%) had participated in research before. For the remaining, the main reason for no participation was not being informed about studies (65%). 172 (43%) preferred regular research updates during a study. The top three ranked research priorities were 1) finding a cure to restore ciliary function; 2) developing treatments to improve lung function and reduce infections and mucus production; 3) finding the best way to manage the disease using existing medication. Other priorities were: involving more doctors and people with PCD in research, raising awareness of the condition, increasing knowledge about mental health and fertility.

**Conclusion:** Our findings will help the PCD research community to improve patient engagement in research and to draw common priorities together with the people who live with PCD and their families.

**Key message:** *What is already known on this topic:*

- In rare diseases, such as primary ciliary dyskinesia (PCD), there are many knowledge gaps, but limited resources, and research priorities should take into consideration topics concerning patients.

*What this study adds:*

- We found that people with PCD and their caregivers are motivated to participate in research when they are informed appropriately and invited, and they would like to receive research updates from studies in which they participate.
- Top ranked research priorities relate to developing new treatments or improving the improved evidence-base for existing treatments to help better manage the condition or to even cure their disease.

*How this study might affect research, practice or policy:*

- By focussing on topics prioritised by people with PCD participation in research may improve.
- Together with already published research priorities of healthcare professionals, our results will contribute to developing a common roadmap for future activities of the PCD research community.

## Introduction

Primary ciliary dyskinesia (PCD) is a rare genetic disease with heterogenous clinical presentation (1). Foremost, PCD affects mucociliary clearance leading to chronic respiratory symptoms and recurrent infections (2–4). Until now, most research was focused on pulmonary disease (5–10) or on improving diagnosis of PCD, which remains challenging (11–14). Generally, PCD research activities were guided by what the research community found interesting and considered a priority, as in most diseases. Engagement of patients in clinical research can ensure that research addresses important and relevant questions and prioritises patient-centred outcomes, leading to greater impact (15).

In recent years, there have been many efforts in different diseases to explore research priorities jointly from the perspective of patients (mostly adults) and researchers across diseases (16,17). The process includes collecting and defining unanswered research questions and prioritising them by importance based on participants’ ranking. In other chronic respiratory diseases, large research networks assessed research priorities of patients and healthcare providers, underlining the value of the patient perspective (18,19).

In the framework of the BEAT-PCD (Better Experimental Approaches to Treat PCD) clinical research collaboration, we explored research barriers and identified priorities in clinical and epidemiological research in the field of PCD from the perspectives of healthcare professionals and researchers (20). This highlighted the need for a dedicated study focused on patients’ and caregivers’ perspectives. We hypothesised that people with PCD might prioritise different research questions compared to healthcare professionals as was in other diseases (21). Therefore, we aimed to explore participation and identify priorities for PCD research from the perspective of patients aged ≥14 years and caregivers of children with PCD.

## Methods

### Study design

We performed a mixed method study consisting of two phases: 1) semi-structured in-depth interviews with people with PCD or caregivers of children with PCD and 2) an anonymous online survey based on results from phase 1. The study received approval from the Bernese cantonal ethics committee (KEK 2020-02250), Switzerland.

### Phase 1: in-depth interviews

We used purposive sampling to select interview participants among people registered in the Swiss PCD registry (CH-PCD) (22). CH-PCD registers all people diagnosed with PCD living in Switzerland. We made sure to include participants from different age groups (adults, adolescents, and caregivers of children) and with varying disease severity to collect rich and diverse data. In addition, we assumed greater motivation and interest in research from participants who had previously participated in CH-PCD surveys and prioritised their invitations. Selected participants received an invitation and study information by post.

Outside of Switzerland, we recruited volunteer participants who responded to our study call circulated by the PCD support groups in Spain and United Kingdom (UK) and through the BEAT-PCD patient advisory group. Volunteers contacted us directly via email and received an invitation with detailed study information afterwards.

In close collaboration with patient partners, representatives from PCD support groups, we developed an interview guide (supplemental document) in German and English for the in-depth, semi-structured interviews. The interview guide was not prescriptive; instead, we encouraged to explore any area and followed new topics opportunistically to ensure depth of information. We conducted and recorded interviews using a teleconferencing software to facilitate notetaking and interview transcription, from March 3^rd^ 2021 to January 17^th^ 2022. Before every interview, we obtained written and verbal informed consent. Participants had the option of having their cameras turned on or off. The first author (YTL) conducted all interviews, after she received training in-depth interviews. Her background as a paediatrician, with experience in talking to patients or parents of affected children, allowed her to notice reluctance or chance for conversational exploration, and relevant behavioural cues.

The purpose of phase 1 was to develop the online survey (phase 2), and the aim was not to achieve data saturation. Therefore, we aimed for a pragmatic sample size of 15–20 interviews, which would still allow to collect rich and detailed data ensuring information power, especially for a rare disease like PCD (23). We transcribed all interviews verbatim focusing on protecting participants’ anonymity by removing identifying information. YTL and last author (MG), female senior researcher, expert in PCD and trained in qualitative research, coded the interviews inductively using a thematic analysis of Braun and Clark approach using NVivo software - release 1.7.1 (24,25). We followed a COREQ (Consolidated criteria for reporting qualitative research) checklist for reporting qualitative research throughout the study (26). Quotes were slightly edited for privacy and clarity and translated by YTL when needed. To ensure anonymity, we present quotes without details on sex or age, just by stating the participant group (adult, adolescent, parent) and a code used for pseudonymisation.

### Phase 2 – Online survey

Based on thematic analysis results of the interviews, we developed an anonymous online survey. Feedback was obtained from patient partners and experts in qualitative research and survey development. We set up the survey online in 8 languages (English, French, German, Greek, Italian, Norwegian, Spanish, Turkish) in a Research Electronic Data Capture (REDCap) study database hosted by the clinical trials unit of the University of Bern (27). We chose the languages based on the participation numbers of people to a previous BEAT-PCD patient survey, choosing the most popular languages (28). We shared the survey link widely via PCD patient support groups worldwide and through the European Lung Foundation. In Switzerland, we had ethical approval to collect personal information and contact participants of CH-PCD for this study. Therefore, we also developed the survey in paper form in German and French and sent it by post to adults, adolescents, and caregivers of children with PCD registered in CH-PCD. Participation was open from June 26^th^ to September 22^nd^ 2023. For ranking questions, we used a 3-, 5- or 7-point Likert scale, ranging from “most important to you” to “least important to you”.

We described characteristics of survey participants using median and interquartile range (IQR) for continuous variables, numbers and proportions for categorical variables. We inquired about and described the median age of the person completing the survey who was either an adolescent or adult with PCD or a caregiver of a child with PCD. We studied possible predictors of staying informed about PCD research and participating in PCD research, including sex, living in a country with a support group, and participation in a support group, using separate multivariable logistic regression models. We ranked research priorities using the mean of a reciprocal ranking score (0–1) to assess the overall top three priorities. Each research question was scored with 1 if ranked first, 1/2 if ranked second, 1/3 if ranked third priority, and 0 if not ranked among the top three priorities. A higher mean score indicated higher priority. We performed analyses using Stata version 15.1.

### Patient and public involvement

Patient partners were involved actively and as co-authors from the conception of the study throughout its whole duration, particularly due to the nature of our research questions. Patient partners as co-authors reviewed the interview guide and adapted it to more patient friendly wording. They also contributed to selecting purposively possible interview candidates among their group members. At phase 2, they reviewed content and wording of questions of the online survey and shared it via their communication channels. We worked and will continue working together with the BEAT-PCD patient advisory group and support groups worldwide to present results of this study in conferences and meetings. We will develop and distribute lay summaries of our findings.

## Results

### Participants

22 people agreed to be interviewed, surpassing our original target. They were 9 adults, 9 parents and 4 adolescents; 16 were female (73%). Median age was 25 years (IQR 27 – 51); 9 lived in Switzerland, 7 in the UK, 3 in other European countries, and 3 in non-European countries.

### Phase 1) in-depth interviews

We identified two broad theme categories from the interviews: developing treatments and increasing knowledge in different fields (figure 1).

**Figure 1:**
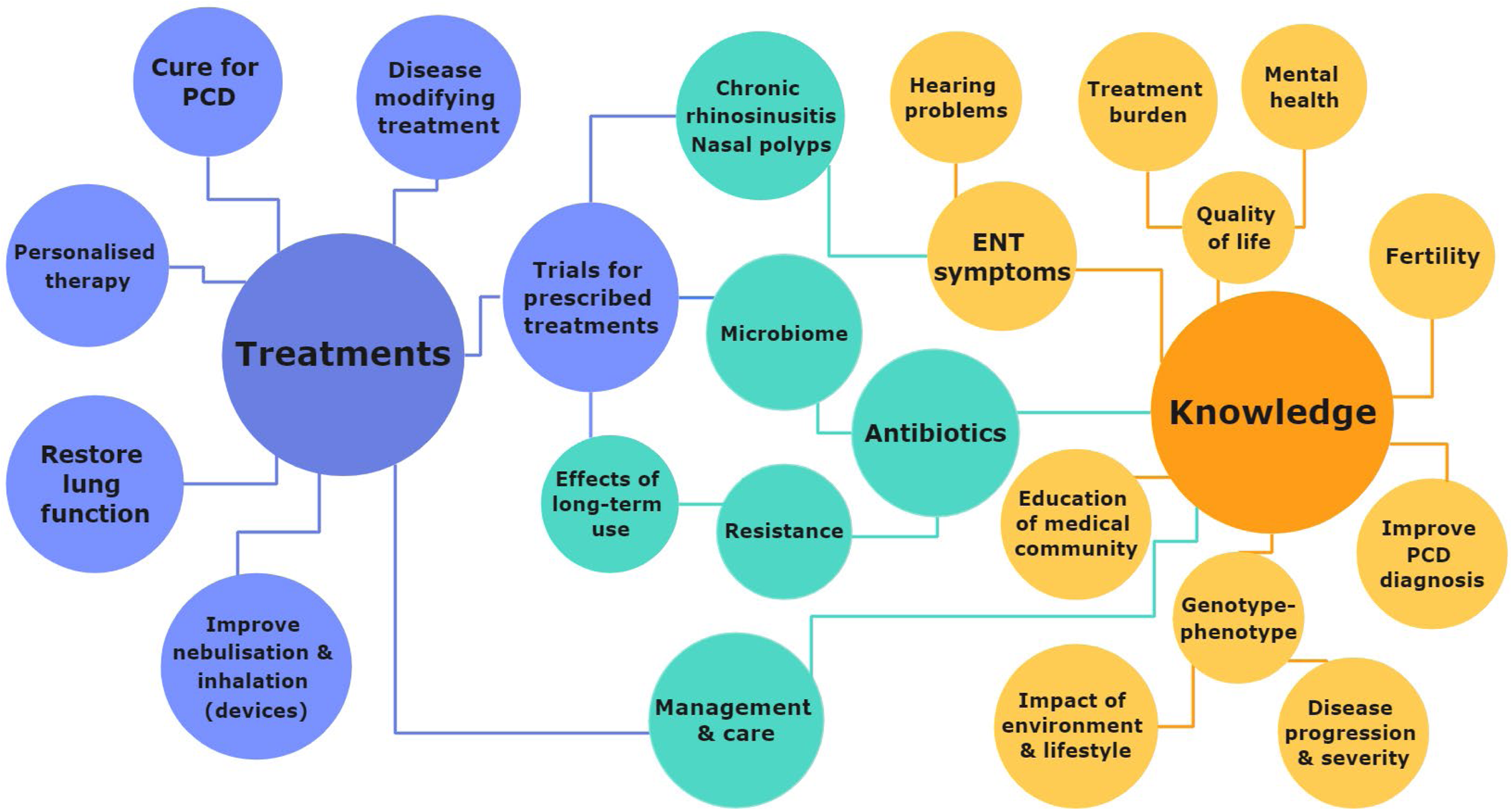
**Main interview themes on research priorities for primary ciliary dyskinesia (PCD) suggested by people with PCD or caregivers of children with PCD. ENT: Ear-nose-throat.**

#### Developing treatments

Developing a cure for PCD or disease modifying treatments, as well as personalised therapy or medications to restore lung function were recurrent themes.

> “What I WANT is a PCD version for the new cystic fibrosis drug that apparently works! That’s what I WANT but that’s very difficult to create I’m going to say. … So if you could make a wonder drug that would be great! But obviously that’s very difficult to do.” (Adolescent; Ea6)
>
> “Obviously, I’m REALLY interested in the possibility of genetic treatments for the condition. Ultimately, this seems like a condition which ought to be curable. […] But maybe one day … maybe by the end of my lifetime will be a point where we will be doing personalised genetic treatments for conditions. I wonder … it would be nice to think we will.” (Adult; EA19)

Concerning treatments, further sub themes were restoring lung function and improving inhalation devices.

Participants mentioned the need for clinical trials for prescribed treatments, questioning the appropriateness of their prescribed treatments, which are often modelled after treatments for cystic fibrosis (CF) without scientific evidence for PCD.

> “And have interventions you have like for … CF! We often model everything on CF, is that appropriate?” (Adult; EA5)

Participants mentioned the effect of treatments on the microbiome and the need for development of new antibiotics. They worried about antibiotic resistance and long-term effects of prescribed treatments.

Developing more effective treatments for nasal polyps was also highlighted.

#### Increasing knowledge

Increasing knowledge in several understudied areas of PCD was another broad theme. Participants requested increasing overall knowledge on ear-nose-throat (ENT) symptoms including hearing problems. When one participant heard about ongoing research on this topic, their reaction was:

> “But that’s fantastic to see that people are more looking into how ENT (symptoms) affect the patients.” (Adult; EA5)

Other themes referred to the need to increase knowledge about the burden of disease as well as the treatment burden, and how they affect mental health and quality of life. Participants felt this topic has been neglected.

> “Everybody with PCD, that I have spoken to, has had a HUGE impact on their ability to work. And yet this is nowhere (mentioned), right? This is not in the literature at all. We talk about disease burden, but you never look UP from the clinic … we never look up really from the patient. […] I feel … that my treatments are a burden, a hindrance. We don’t look at the lived ramifications of that, which is that many people are NOT working full time because of PCD. They’re STRUGGLING to work. Whether young or old … it seems to be a theme. I just don’t think it’s come up yet.” (Adult; EA9)

Participants wished for studies to increase knowledge on fertility in PCD as well as improving PCD diagnosis. They also asked for better understanding of genotype-phenotype correlations, their associations with disease progression and the potential impacts of environmental factors and lifestyle choices.

Another important theme was increasing the awareness of PCD in the medical community especially of general awareness for PCD, as patients or their family members often had to explain the disease to their doctors.

> “Because in the past, my mother took us to the emergency room, and she had to explain to the doctors what PCD was. Having to go through this process is NOT okay. I think the doctors MUST know what PCD is. I know it’s a rare disease and it’s not common. But this is not a reason for not knowing PCD. Doctors must know what PCD is, just as they know other diseases like cystic fibrosis. But nobody knows PCD. So we have to inform doctors about PCD ourselves.” (Adolescent; Ea17)

An overarching theme related to both broad categories was improving PCD management and care by establishing specialised PCD care centres. We included a detailed list of themes and specific questions relating to each theme in the supplementary section under (table S2).

#### Phase 2) Online survey

399 participants completed the survey with median age 41 years (IQR 33–49), living in 29 different countries; 74% were female (table 1). Half (52%) had PCD themselves, while the others were caregivers of a child with PCD. 286 (72%) participants knew there was a PCD support group in their country of residence (supplemental table S1). Among them, only half (51%) participated in activities or were representatives of the group. 86 (23%) participants living in countries with a national PCD support group did not know about its existence.

**Table 1:** Characteristics of survey participants (N=399)

|  | N (%) |
| --- | --- |
| **Country of residence** |  |
| France | 70 (17.5) |
| Germany | 64 (16) |
| United States | 42 (10.5) |
| Switzerland | 40 (10) |
| Italy | 38 (9.5) |
| United Kingdom | 23 (6) |
| Spain | 20 (5) |
| Australia | 11 (3) |
| Other European countries | 69 (17) |
| Other non-European countries | 12 (3) |
| Not reported | 10 (2.5) |
| **Sex** |  |
| Female | 295 (74) |
| Male | 100 (25) |
| Not reported | 4 (1) |
| **Age of participants in median years (IQR)<sup>a</sup>** | 41 (33 – 49) |
| **Connection to PCD** |  |
| Person with PCD | 207 (52) |
| Caregiver of child with PCD | 167 (42) |
| Not reported | 25 (6) |

PCD: primary ciliary dyskinesia. Characteristics presented as N (%) or median (interquartile range (IQR)) unless otherwise stated. <sup>a</sup>Among 380 participants (person with PCD and caregivers of children with PCD) who reported their age. Age represents the age of the person completing the survey, not the person living with PCD.

Two thirds (67%) of participants said they remained informed about research related to PCD, mostly through word of mouth (42%) (table 2). About half (45%) had participated in PCD research before, of them 35% were invited by their physician to participate. The reasons for not having participated in PCD research before were most commonly not being informed about studies (65% of 168 participants) or not fitting the inclusion criteria (8%). Other reasons were that studies took place in different countries or more than 10 driving hours away, because they were only recently diagnosed, or there had not been any opportunities yet.

**Table 2:**
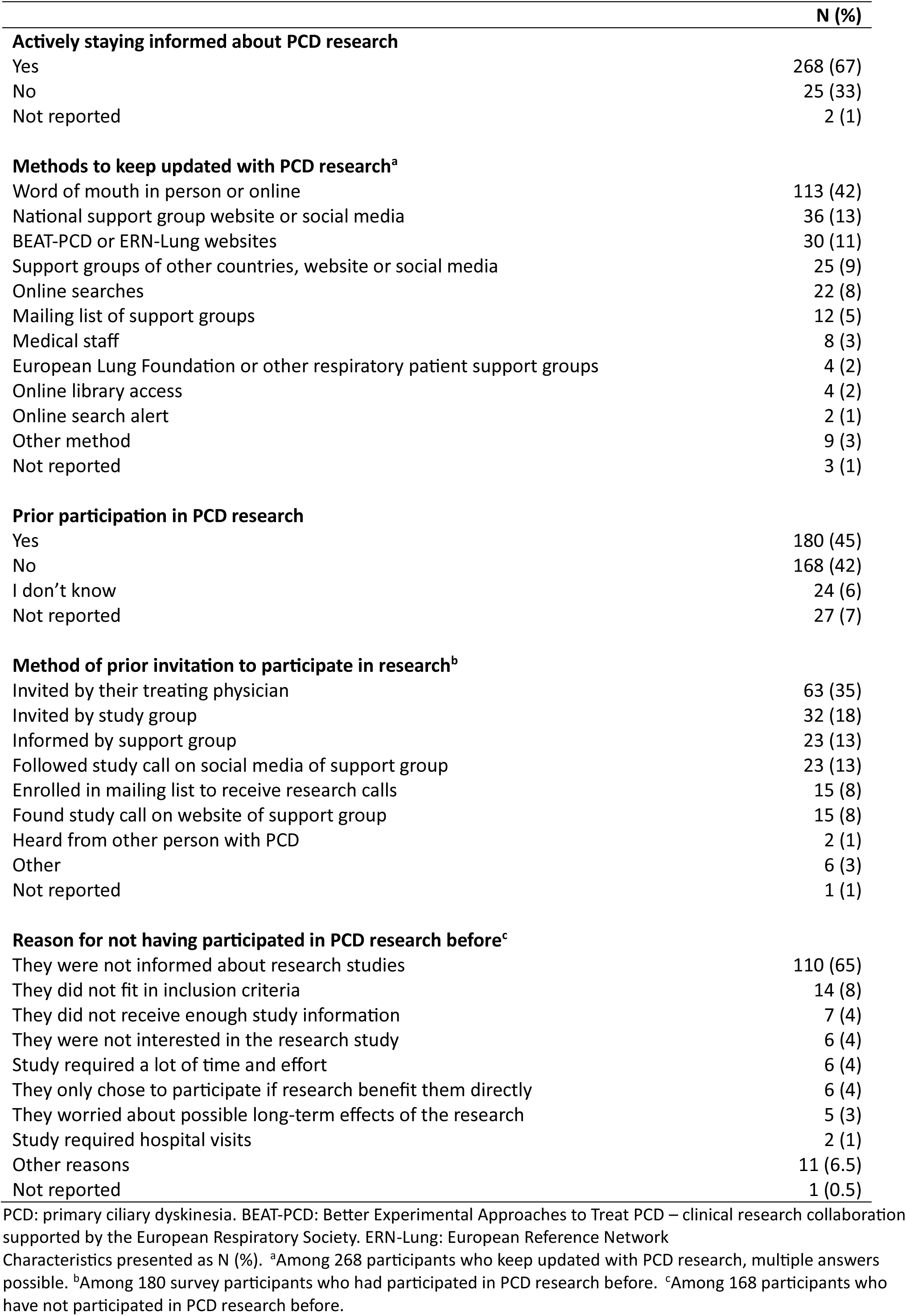
Experiences from previous participation in PCD research among survey participants (N=399)

Participation in a national support group appears to have a positive effect on both in staying informed about PCD research (OR 1.4, 95% CI 0.9 – 2.3; supplemental figure 1a), and on participating in PCD research (OR 1.5, 95% CI 1.0 – 2.4; supplemental figure 1b). We did not find any association of staying informed about or participating in PCD research (supplemental figure 1b) with being a person with PCD or a caregiver, man or woman, or living in a country with a PCD support group.

Among the 180 participants who had participated in PCD research before, about half (46%) had received study results (table 3). When received, results were understandable for most people. Participants preferred receiving regular updates by email or on a study website during the study duration (43%). Only 2% of participants were not interested in receiving any results. Participants preferred receiving feedback during the study (52%) compared to asking back about results (32%).

**Table 3:** Experiences and preferences about receiving feedback on PCD research results among survey participants (n=399)

|  | N (%) |
| --- | --- |
| **Received study results after study completion<sup>c</sup>** |  |
| Yes, received results/updates without any efforts | 70 (39) |
| Yes, but had to ask for results | 13 (7) |
| No, but research is still ongoing | 40 (22) |
| No, never heard back | 36 (20) |
| Not sure | 17 (10) |
| Not reported | 4 (2) |
| **If research results were received<sup>b</sup>** |  |
| They understood all information | 53 (64) |
| They understood some parts | 26 (31) |
| They did not understand most of the information | 1 (1) |
| They do not remember | 3 (4) |
| **Preferred time to receive research results** |  |
| Receive regular updates and results during the study via email or on website | 172 (43) |
| Receive final study results at the end of the study | 88 (22) |
| Receive regular updates and results during the study online or on-site with research team | 80 (20) |
| Not interested in receiving research results | 8 (2) |
| Not reported | 51 (13) |
| **Preferred method to receive research results<sup>c</sup>** |  |
| Study website | 97 (24) |
| Email | 75 (19) |
| Video | 17 (4) |
| Letter | 16 (4) |
| Lay summary | 15 (4) |
| Scientific publication | 5 (1) |
| Own result in addition | 10 (3) |
| Not reported | 164 (41) |
| **Preferred to** |  |
| Receive feedback during research without actively requesting | 207 (52) |
| Ask actively about research results when they wish | 127 (32) |
| Not reported | 65 (16) |

PCD: primary ciliary dyskinesia. Characteristics presented as N (%). <sup>a</sup>Among 180 survey participants who had participated in PCD research before. <sup>b</sup>Among 83 participants who had received research results. <sup>c</sup>Multiple replies possible.

Among the research priorities participants were asked to rank, the top three priorities were: 1) “Can we find new medication that will ‘cure’ PCD or reduce the need for treatment by restoring the function of cilia in the body (like the medication available in cystic fibrosis)”; 2) “Are there treatments that will improve lung function, reduce infections, and reduce the amount of mucus I produce?”; and 3) “What is the best way to treat PCD (including lungs, ears, and nose) using existing medication and other management approaches?” (table 4). Ranking scores on overall top priorities varied, ranging from 0.04 to 0.48, highlighting that many topics were ranked highly by participants even if not included in the top of the list. Comparing research priorities by sex, men ranked research on life expectancy as more important and research on ENT symptoms as less important, compared to women. In the final question, participants could suggest other research topics they considered a priority that were not already listed. Answers included: having genetic counselling for family planning and foetal genetic testing, defining which specific exercises can be done at home to manage one’s symptoms, and studying the effects of PCD on the sense of smell.

**Table 4:** Overall top research priorities for primary ciliary dyskinesia (PCD) as ranked by people with PCD or parents/carers of children with PCD, overall and by sex of survey participants.

| Rank | Research topics | Reciprocal mean score |  |  |
| --- | --- | --- | --- | --- |
|  |  | Total<br>(n=399) | Females<br>(n=295) | Males<br>(n=100) |
| 1 | Can we find new medication that will 'cure' PCD or reduce the need for treatment by restoring the function of cilia in the body (like the medication available in cystic fibrosis) | 0.479 | 0.474 | 0.483 |
| 2 | Are there treatments that will improve lung function, reduce infections, and reduce the amount of mucus I produce? | 0.220 | 0.209 | 0.252 |
| 3 | What is the best way to treat PCD (including lungs, ears, and nose) using existing medication and other management approaches? | 0.204 | 0.192 | 0.213 |
| 4 | Can we find new antibiotics or other medication to tackle antibiotic resistance, and what are the effects of long-term antibiotic use? | 0.129 | 0.135 | 0.110 |
| 5 | How can we get more doctors and people with PCD involved in research and make them aware of this condition? | 0.127 | 0.125 | 0.130 |
| 6 | What health-related behaviours can I do, and what everyday things should I avoid to control the improvement or worsening of my symptoms and quality of life? | 0.094 | 0.077 | 0.133 |
| 7 | How is mental health affected in people with PCD and their families (psychological aspects of PCD e.g. treatment burden and coping in daily life)? | 0.074 | 0.082 | 0.053 |
| 8 | What is the life expectancy in PCD and what are the long-term impacts of this disease and its treatments? | 0.056 | 0.051 | 0.073 |
| 9 | How is fertility affected in patients with PCD and what are the best fertility management approaches? | 0.055 | 0.059 | 0.050 |
| 10 | What is the best medication plan for each patient with PCD, including specific dosages and route (orally or intravenous)? | 0.054 | 0.057 | 0.048 |
| 11 | How should PCD be managed correctly in different age groups (also in people without symptoms)? | 0.051 | 0.052 | 0.045 |
| 12 | How does PCD affect the ears, balance issues, nose, and sinuses and how are these problems linked with problems in the lungs? | 0.049 | 0.059 | 0.020 |
| 13 | How does PCD affect the gastrointestinal system, such as stomach and intestines (e.g. reflux, indigestion, bloating)? | 0.047 | 0.052 | 0.030 |
| 14 | Are specific genes associated with specific symptoms or more severe disease (genotype-phenotype correlation)? | 0.044 | 0.051 | 0.025 |
| 15 | How can the treatment duration and treatment efforts be reduced without compromising effectiveness? | 0.039 | 0.040 | 0.038 |

Research topics ranked from most to least important (listed among top three overall priorities) based on the mean of reciprocal ranking score (0–1); each question was scored with 1 if ranked first, 1/2 if ranked second, 1/3 if ranked third, and 0 if not ranked among the top three priorities. PCD: primary ciliary dyskinesia. 4 participants preferred not reporting their sex and were excluded from the stratified scores

## Discussion

With a mixed-method design, we assessed participation of people with PCD and caregivers of children with PCD in research and identified research priorities from their perspectives. Our findings inform the PCD research community about opinions and preferences of patients regarding their participation in research and the communication of results. The study also highlights the importance of PCD support groups, which should be supported by healthcare professionals. Close collaboration of researchers with support groups will encourage further participation of people with PCD in upcoming research and facilitate wider dissemination of results in an appropriate format. The results of this study, taken together with the priorities identified by PCD professionals and researchers (20), will strengthen future research, especially of large research networks, such as BEAT-PCD, and contribute that it does focus on questions most relevant to PCD patients and their families.

Ours is the first study to explore participation in research and research priorities for PCD from the perspective of patients as well as adolescent PCD patients and caregivers of children with PCD. Performing semi-structured interviews allowed us to explore patients’ perspectives in-depth in order and to develop a survey that included many different opinions. Our study was strengthened by the involvement of patient partners in all study steps, from the very beginning with the development of the interview guide to creating the survey. The majority of survey participants were female, which may have led to underrepresentation of male participants, a common phenomenon in online surveys (28). The survey was distributed during summer, which might have affected participation, however the 3-month duration ensured any such effect was minimal. Our survey was circulated worldwide and was available in 8 languages ensuring wide reach (399 participants from 29 countries), but our study call reached only people with connections to PCD support groups, BEAT-PCD, or the European Lung Foundation. People outside of this circle, without knowledge of the survey languages, or who had no ability to participate online could not participate.

We found collaboration between researchers and patient support groups to be an important factor that can contribute to research participation, which was also described in a systematic review of approaches for engaging patients for research on rare diseases (29). Preferably, participants would like to receive research updates during the study process and most importantly receive any feedback in an appropriate manner. We found at least one third of participants had not received any results from studies in which they had participated. If received, participants stated that results were mostly understandable, but this could further be improved with use of lay summaries co-developed with patient partners. Reporting results back in lay summaries is a growing effort among researchers to convey their results directly without misconstruction or misinterpretation (30). Preferred methods of receiving research results or feedback was digital media, which facilitates active real-time public communication and research visibility benefitting future research participation (31). In a qualitative study with semi-structured interviews with research participants and patient advisors about contributing to health research, the authors reported that communication between research teams, participants, and clinicians could be improved e.g. by simplifying study documentation and providing feedback on findings (32). We also found this theme in our interviews along with improving knowledge and awareness of PCD in the medical community.

The highest ranked research priority was finding a cure for PCD; all top three ranked priorities were related to treatments to manage symptoms and improve quality of life. While these results are not surprising they highlight the need for further clinical trials on the management of PCD as current evidence is limited (33–35). This study also confirms the enthusiasm and anticipation of people with PCD regarding the new potential molecular treatments that are in the pipeline. Interestingly, ranking varied a lot, underlining that participants also prioritised other questions. It is possible that they prioritized more realistic aims or short-term priorities that could be achieved in the near future. We unfortunately did not differentiate between short-term and long-term priorities in our survey.

Compared to healthcare professionals, patients with PCD and their caregivers identified many similar priorities especially related to management of the disease and its symptoms (20). However, the ranking of priorities often differed e.g. mental health was ranked much higher by patients than professionals. Topics raised exclusively by patients were related to effects of medication and treatment burden e.g. antibiotic resistance, long-term impacts of PCD treatments, and how treatment efforts can be reduced without compromising effectiveness. Several areas that were identified to need further research such as treatment burden, fertility, ENT disease, and genotype-phenotype correlations are topics of recent or planned studies, some of which with extensive patient involvement, which shows that the PCD research community considers more and more the perspective of people with PCD and their families (36–38,2,3,39).

Our findings of patient preferences on participation in research and priorities for PCD research contribute to improvement of future recruitment strategies and communication of research in an appropriate manner for the PCD patient community. This will further encourage participation in future research. Jointly, priorities identified by people with PCD and PCD professionals, and continued collaboration with patient partners will undoubtedly help to plan more successful future research activities within the field of PCD.

## Data Availability

The datasets used and analysed during this study are available from the study PI Dr Myrofora Goutaki upon reasonable request.

## Acknowledgements

We thank all people with PCD and their families participating in the study (especially PCD Support UK; Kartagener Syndrom und Primäre Ciliäre Dyskinesie e. V. Deutschland/ Deutschschweiz; Association ADCP France; RespiFIL; Asociación Nacional de Pacientes con Discinesia Ciliar Primaria DCP España/PCD Spain; ePAG Georgia; The Organisation for Respiratory Health in Finland (Hengitysliittory); Swedish Primary Ciliary Dyskinesia Support Group; Associazione italiana discinesia ciliare primaria - sindrome die Kartagener Aps; PCD Supportprimer silyer diskinezi derneǧi (SiLYADER); PCD Foundation) and the European Lung Foundation for their collaboration. We thank Daniela Knup (University of Bern) for her assistance with the transcription of some interviews.

## Author contribution

M. Goutaki developed the concept and design of the study. Y.T. Lam managed the study under supervision of M Goutaki. Y.T. Lam conducted and transcribed the in-depth interviews. Y.T. Lam performed thematical analysis, cleaned and standardised data, performed statistical analysis and drafted the manuscript supervised by M. Goutaki. All authors contributed to development of the interview guide and the online survey and commented on the manuscript.

