## Supplemental material for "Patients’ research priorities and participation in primary ciliary dyskinesia research"

**Table S1: Participation in and knowledge of PCD patient support groups among survey participants (N=399)**

| Is there a patient support group in your country and are you involved in it? | N (%) Total | N (%) Countries with patient group[a] | N (%) Countries without patient group[b] |
|---|---|---|---|
| Yes, there is a patient group. I do not participate in it. | 141 (35) | 139 (38) | 2 (5) |
| Yes, there is a patient group. I attend meetings and participate in activities. | 107 (27) | 101 (28) | 6 (17) |
| Yes, there is a patient group. I am a representative/ committee member. | 38 (10) | 37 (11) | 1 (3) |
| No, there is no patient group. | 48 (12) | 24 (6) | 24 (67) |
| I don't know if there is a patient group. | 65 (16) | 62 (17) | 3 (8) |

PCD: primary ciliary dyskinesia. Characteristics presented as N (%). [a]Among 363 participants living in countries with known PCD support groups. [b]Among 36 participants living in countries which do not have PCD support groups.

**Table S2:**

**Themes and specific questions related to research priorities for primary ciliary dyskinesia (PCD) from in-depth, semi-structured interviews with people with PCD or caregivers of children with PCD**

**Treatment strategies**

1. Can we find a cure for PCD?
2. Can we test if CTFR modulator therapies work in PCD?
3. How can we improve ciliary beating everywhere in the body?
4. Can we test if Pulmozyme helps in PCD?
5. Can we test medications which are prescribed for PCD?
6. Can we test and compare inhalation devices for PCD?
7. Can we develop a physiotherapy app for PCD?
8. Can we develop a drug to reduce mucus production in PCD?
9. Can we develop a treatment for nasal polyps in PCD?
10. What are long-term treatment effects in PCD?
11. Can we develop individualised treatments by dosage and with different application routes for PCD?
12. Can we develop treatments to improve quality of life in PCD?
13. Can we stabilise lung function and reduce infections in PCD?
14. Can we develop new antibiotics?
15. Can we develop other new antimicrobic treatments to tackle antibiotic resistance?
16. When are intravenous or oral antibiotics more beneficial in PCD?
17. Can we test bronchiectasis treatments for PCD to reduce/reverse lung damage as tested in COVID-19 patients e.g. re-transfusion of selected white blood cells?

**Symptoms**

18. Can we study all symptoms related to PCD and look into differences by sex in PCD?
19. Can we study ENT symptoms and hearing problems in PCD?
20. Can we study gastrointestinal problems in PCD?
21. How is the genotype-phenotype correlation in PCD?
22. What are long-term disease effects in PCD?

**Upper airways**

23. Can we study effects of chronic rhinosinusitis on sinus surgery in PCD?
24. What is the benefit of sinus fenestration in PCD?
25. Can we develop hearing tests for home use to adjust hearing aids in PCD?
26. Can we study balance disorders in PCD?

**Microbiology**

27. What is the importance of certain pathogens in PCD?
28. What are long-term effects of taking antibiotics (e.g. on the gastrointestinal tract) in PCD?

**Health-related behaviours/mental health**

29. Can we study if sports or saline inhalation are better in mucus clearance in PCD?
30. Can we test if homeopathy or sauna reduce symptoms in PCD?
31. Can we study nutritional effects on symptoms in PCD in PCD?

32. Can we study how treatment duration affects daily life?
33. Can we research treatment burden in PCD?
34. Can we study psychological, psychosocial aspects and needed support in PCD?
35. How do people with PCD cope in daily life?
36. Can we study how nutrition and minimally needed fluid intake affect symptoms in PCD?

**Comorbidities**

37. Can we improve understanding of PCD to differentiate between other diseases?
38. Are learning difficulties associated with PCD?
39. Are dental abnormalities e.g. enamel issues associated with PCD?

**Diagnosis**

40. How can we improve diagnosis in PCD?

**Research on special groups**

41. How can we include more research including adults in PCD?

**Management and care of PCD**

42. Can we build PCD centre (for adults)?
43. How do we improve transition to adult care in PCD?
44. How can we increase PCD knowledge and management among medical staff to improve better treatment outcomes?
45. How can we improve multidisciplinary communication and treatment e.g. integration of treatment/management, social aspects in daily life in PCD?

**Other**

46. How is fertility affected and what are the best fertility management approaches in PCD?
47. Can we develop information (booklet) about PCD (what is PCD, how is it managed, what is the prognosis) – to have a clear guide to give to people or parents at diagnosis?
48. What causes fatigue in PCD and how can we treat it?
49. Can we study pain perception in PCD?

PCD: primary ciliary dyskinesia. CTFR: cystic fibrosis transmembrane conductance regulator. COVID: Coronavirus disease 2019

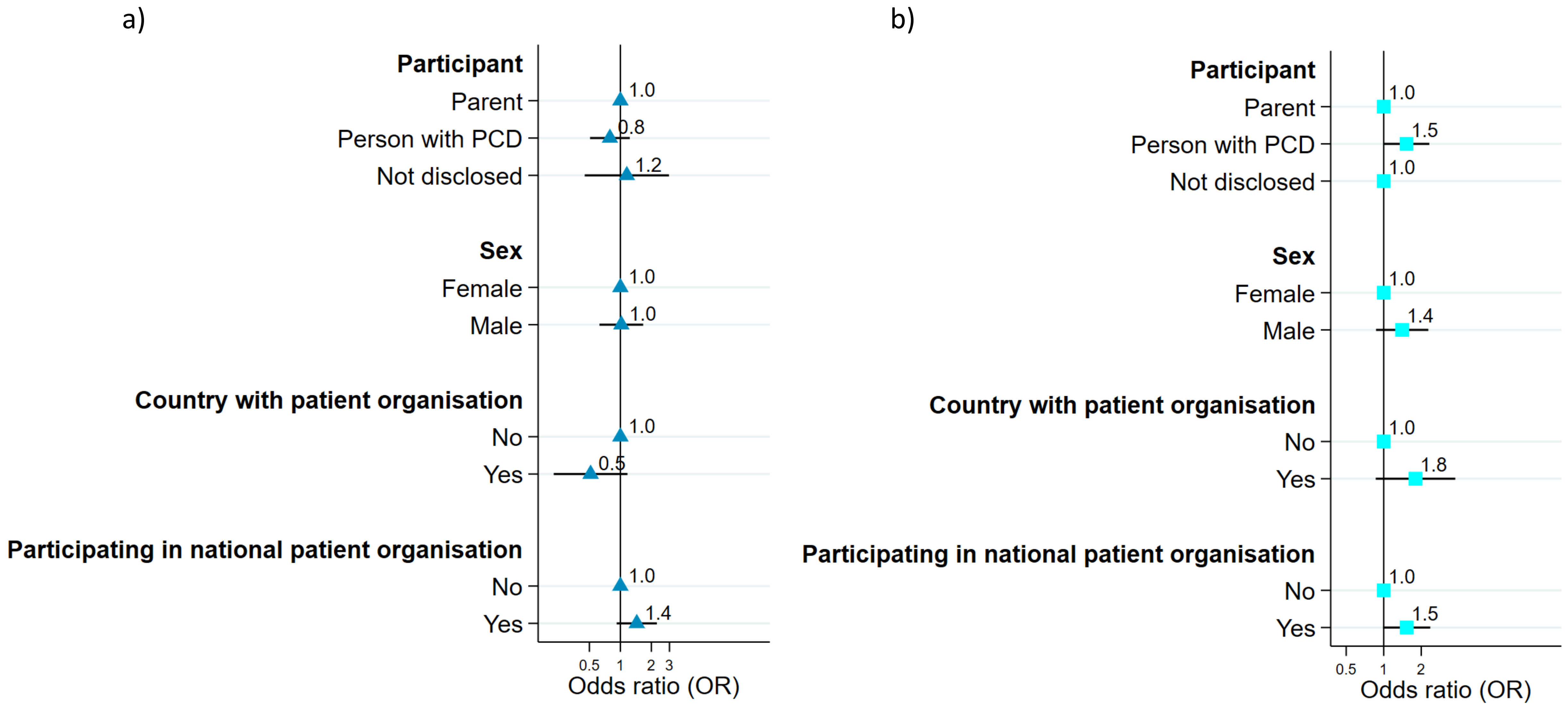


**Figure S1:** Participation of survey participants in research related to primary ciliary dyskinesia (PCD) N=399
a) Staying informed with PCD research b) Participation in PCD research
